# Comparing Classifier Performance to Predict Infectious Diseases

**DOI:** 10.1101/2023.05.06.23289606

**Authors:** Roger Geertz Gonzalez

## Abstract

We compared the accuracy of the machine learning classifier algorithms: Random Forest, Naïve Bayes, Decision Tree, and Artificial Neural Network to predict zoonoses using the Random Forest extracted features and the serology data for seven different zoonotic diseases as the targets. We identified Random Forest and Naïve Bayes as having the best performance overall. The Random Forest models above did well using Positive Predictive Value (PPV), Area Under the Curve (AOC) and Receiver Operating Characteristic (ROC) performance measures in identifying the positive cases for each of the diseases which is imperative when it comes to being able to identify the disease and then use this information to implement prevention and medical aid to specific areas and people where it is most needed. It also does well in predicting the negative values which is important to ensure the negatives are not false negatives.

Naïve Bayes was found to be the best choice for accuracy and performance. NB works well because it treats each feature as independent and thus, any change in one feature will not affect the other in the NB model. Decision Tree could not capture the data and thus, underfit during the first initial modeling and after hyper tuning. Artificial Neural Network overfit the model by capturing all the data including noise in the initial model, but underfit after hyper tuning. Both Decision Tree and Artificial Neural Network classifier algorithms are not recommended as classifiers for this dataset.

**Statements:** There are no conflicts of interest in this work.

All methods were carried out in accordance with relevant guidelines and regulations.

All experimental protocols were approved by the Forestry Administration of Cambodia.

Informed consent was obtained from all subjects and/or their legal guardian(s) at the beginning of the survey.

## 1. Introduction

KAP studies have mainly used regression models to study the relationships between zoonoses and social attitudes. For example, Kiffner et al. (2019) use linear mixed models to study the relationships between anthrax rates and human attitudes and practices in Tanzania. In their study of rabies in Bhutan, Rinchen et al. (2019) use multivariable logistic regression to estimate positivity rates. Saylors et al. (2021) similarly used multivariable logistic regression to study zoonoses and wildlife practices trade in Cameroon. Head et al. (2020) used logistic regression to examine zoonotic risk factors via serology and practices and beliefs related to Crimean-Congo Hemorrhagic Fever in Kazakhstan. However, the major limitations to these studies that use linear and logistic regression for statistical inference modeling is that they assume a linear relationship between the dependent variable and the independent variables (Lantz, 2019). In complex real-life situations relating to infectious diseases including causes, effects, and transmission, the relationship between the dependent variable and independent variables might be non-linear and not an oversimplified linear or logistic regression model. Another limitation for linear and logistic regression is that the independent variables (features in machine learning) must be known ahead of time and tested and then retested to ensure the model works. Additionally, statistical inference models usually use conservative analysis strategies, but methods in machine learning are more flexible (Yoo et al., 2012). Lastly, while statistical inference models build hypothesis and then use collected data to test the hypothesis, machine learning can explore hidden patterns from collected data without a hypothesis.

Machine learning algorithms can replace both linear and logistic regression for both regression and classification problems for complex, real-life problems such as disease prediction and features do not need to be known ahead of time and then retested to ensure the best fitting model. For example, RF has been used successfully in disease prediction when it comes to identifying important features.

Velusamy & Ramasamy, (2021) used the Random Forest (RF) algorithm successfully to select important features for their combined K-Nearest Neighbor, RF, Support Vector Machine (SVM) algorithm classifiers to predict coronary heart disease using the Z-Alizadeh Sani medical dataset. They found that a Boruta based RF algorithm combined with a SVM feature extraction was the best combination that had the best predictive results. Zhao et al. (2020) were also able to successfully incorporate RF as a dengue forecasting tool in their Colombia research. Yadav & Pal (2020) also use RF to identify the most important features that predict heart disease. They found their RF to be 99% accurate. Alam et al. (2019) found that RF was highly accurate predicting different diseases from 10 different disease datasets (breast cancer, diabetes, bupa, hepatitis, heart-statlog [heart disease], SpectF [heart disease], SaHeart [heart disease], PlanningRelax [EEG tests for stress], Parkinsons, and hepatocellular carcinoma).

In their study predicting cancer, Uddin et al. (2019), found that RF, Naïve Bayes (NB), Decision Tree (DT), and Artificial Neural Network (ANN) were highly accurate machine learning classifiers using sensitivity, specificity, and ROC/AUC rates. Alshereff et al. (2019) found that RF, DT, and ANN were highly accurate in predicting blood diseases. Fatima & Pasha (2017) similarly found that DT and ANN were able to highly predict dengue disease.

Although, these studies show how these machine learning models can be accurate, some machine models can overfit (good performance on training data, but poor generalization to test and other data) or underfit (poor performance on training data and poor performance on generalizing to test and other data) the data. In these instances, hyper tuning of parameters is necessary to ensure the model is accurate (Lantz, 2019).

The objective of this study is to compare machine learning classifiers and then see which one works best to predict zoonotic diseases from the KAP survey and serological data. Because of its high prediction rates, we used RF feature extraction to determine the most important features in the KAP survey. These features were used in the RF, NB, DT, and ANN classifiers to compare their predictive results regarding the KAP survey and the blood virus antibody tests since these classifiers have been shown to be predictive in other health fields.

Some specific advantages to RF include: a. it reduces overfitting that usually occurs in DT and improve accuracy, b. it works well with both continuous and categorical variables, c. takes into account non-linear effects, d. normalizing data is unnecessary because of its rule-based approach, and e. it can identify important features (Lantz, 2019). The major disadvantage of RF is that it requires lots of computational power to create numerous trees before providing the specific output. Some advantages to using NB include: a. the algorithm works quickly and saves time and b. since is assumes all features are independent, it performs better than other models and requires less training data. Some disadvantages using NB are: a. the assumption of all features being independent does not happen frequently in real life and b. its estimations can be wrong sometimes (Lantz, 2019). Some advantages to using DT include: a. requires less effort for data preparation, b. normalization and scaling of the data are not required, c. its rule-based techniques are very easy to explain and interpret. Some disadvantages to using DT include: a. small change in the data can cause instability when creating trees and b. it takes time to train the model (Lantz, 2019). The advantages to using NN include: a. can be used for complex, non-linear problems, b. it can overfit, and c. a failure in some of its nodes does not prevent it from producing an output. Its disadvantages can include: a. its “black box” design can prevent interpretability of results and b. appropriate network structure is achieved by trial and error which is time consuming (Lantz, 2019).

Infectious disease modeling is essential for understanding and testing different public health strategies to prevent future epidemic outbreaks (Dattner & Huppert, 2018). Unlike KAP studies that use either linear or logistic regression for inference, or to determine the relationship between variables in their studies of infectious disease (Funk & King, 2020), this study uses machine learning prediction techniques to predict positive and negative cases and to determine which features are the most important that might cause positive cases. There are recent examples where machine learning methods to predict infectious diseases were successful. For example, Han et al. (2015) used machine learning techniques to identify reservoir status with high accuracy and predicted new hyperreservoir (harboring 2 or more zoonotic pathogens). Colubri et al. (2016) created a machine learning model to predict clinical outcomes in patients seropositive with Ebolavirus during the 2013-2016 West African epidemic.

## 2. Material and Methods

The KAP dataset initially included 1656 instances (participants) and 375 features of combined survey and serology data. Questions that were mostly left blank were deleted from the dataset which included many participants that did not answer most questions. This left 896 instances and 105 features (Table 2.1). Seropositivity cutoffs were developed using 3-fold change above the arithmetic mean of the mock-adjusted scaled MFI data (shown as the solid straight line in the figure below) as well as fitted to a log-normal model (shown by dashed lines below) (Colubri et al., 2016) (Figure 2.1). Viruses that exceeded the highest threshold, the 3-fold change above the arithmetic mean was considered seropositive (MENV, BOMBV, EBOV, BDBV, TAFV, SUDV, RAVV, LLOV, MLAV, MOJV, HEV, CEDV, and GHV).

**Table 2.1.**
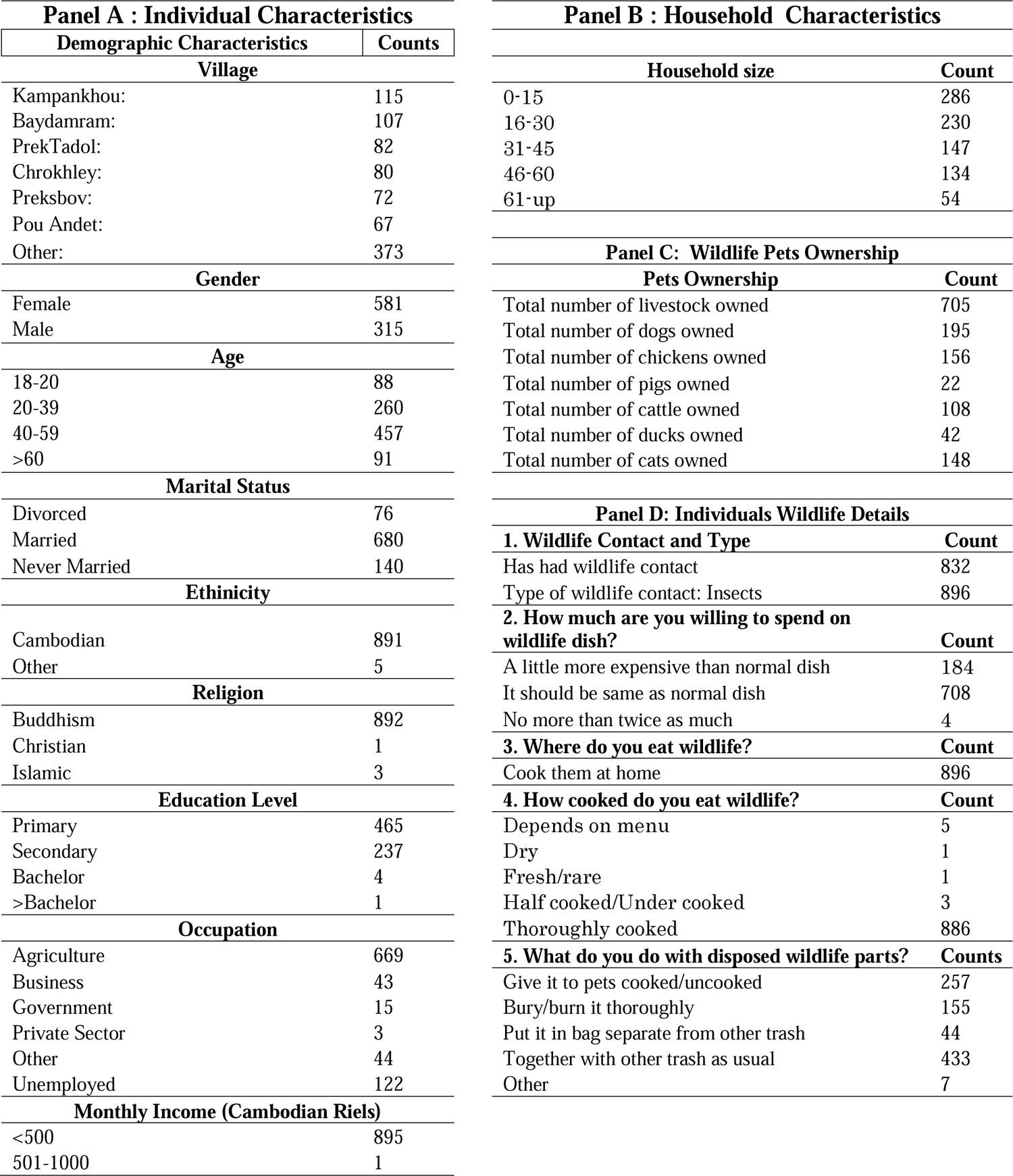
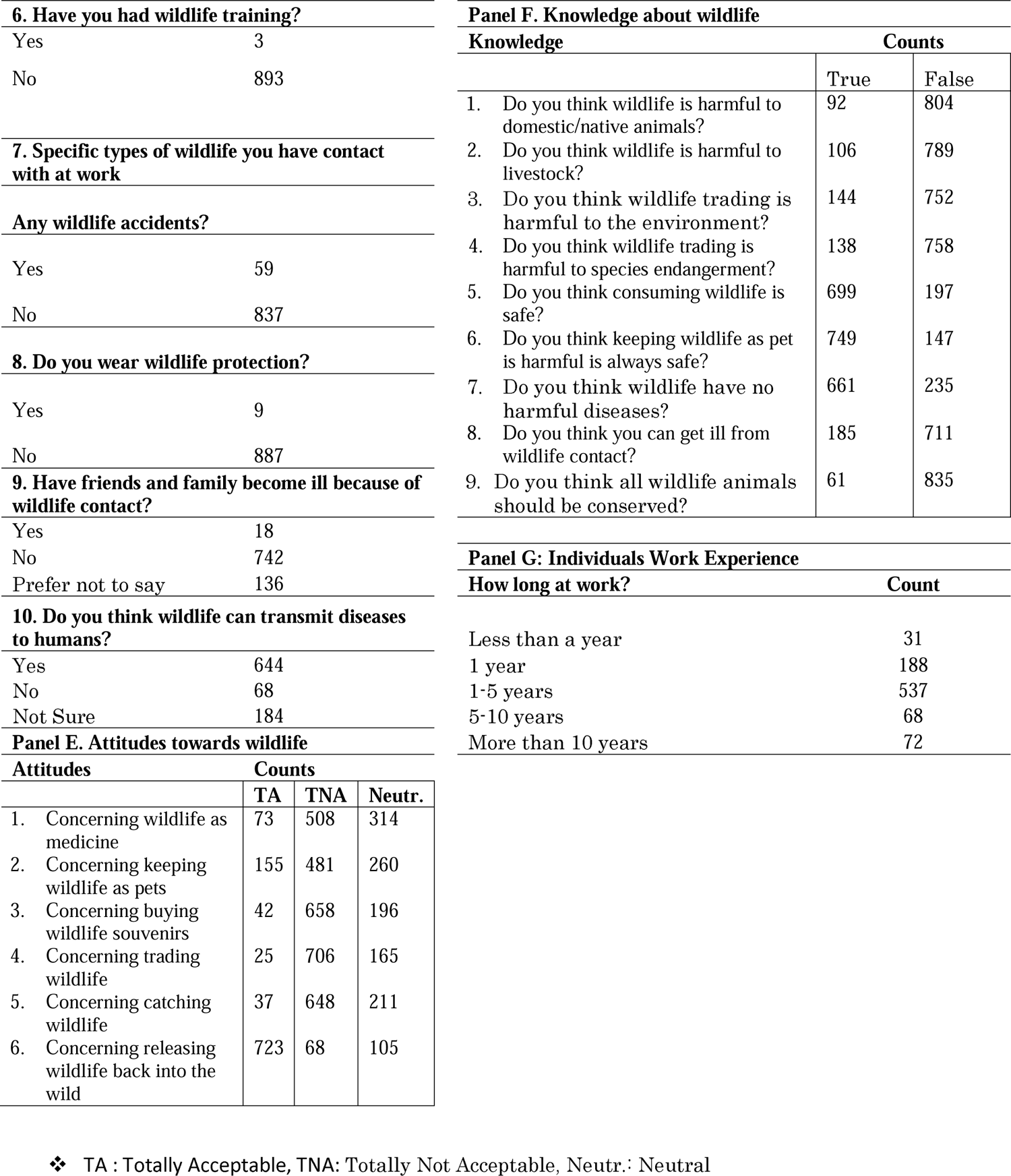
Feature Counts Used for Random Forests Classifier and Feature Extraction

**Figure 2.1.**
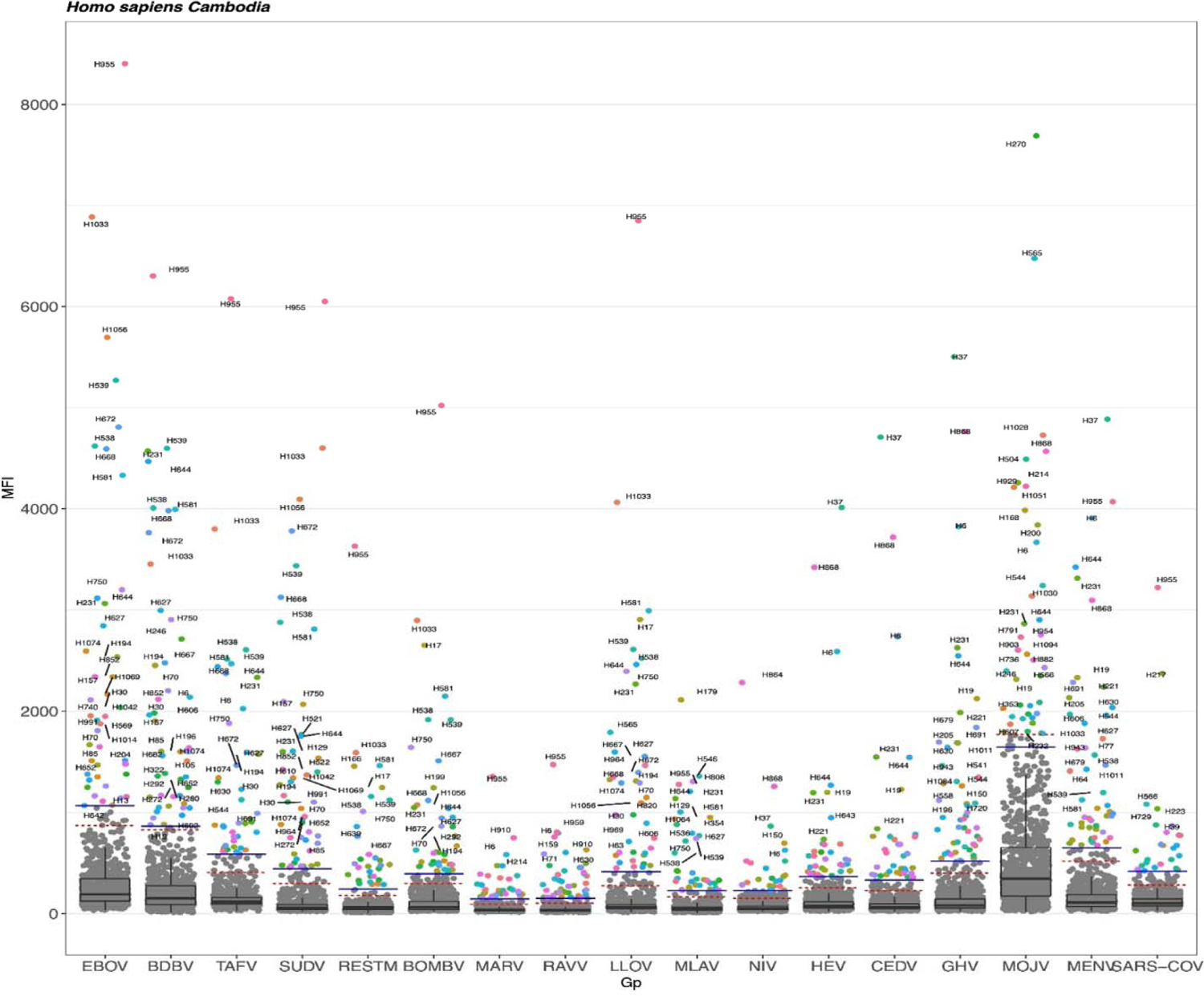
MFI Cutoffs

Analyses for this study was done in R, version 4.1.0 (R Core Team, 2020). MFI 3 times above the arithmetic mean cutoffs are designated by a solid horizontal line (Figure 2.1). A dashed line indicates the log-normal cutoff. The 3 times the mean arithmetic cutoff was the highest positivity threshold used for this study. Survey answers had a range of missing values between 0% to 95%. Missing values accounted for 9.81% of the data and these were all replaced with imputation. Missing imputation (MI) can reduce bias and improve efficiency for analysis of Missing at Random (MAR) data at any proportion of missingness (Madley-Dowd et al., 2019). *missForest* was used as an imputation-based method on the Random Forests (Stekhoven & Bühlmann, 2012). It uses iterative imputation by training an RF in observed values followed by predicting the missing values and then proceeding iteratively. It works well with both continuous and categorical data, and it takes into account non-linear effects (Stekhoven & Bühlmann, 2012). *missForest* iterated for a total of 9 times and replaced the missing values.

Data that show 0% to 4.7% positivity disease rates are difficult when creating training and testing for classifiers assume that the test data is from the same distribution as the training data (Khalilia et al., 2011). For this study, only diseases that had at least 4.7% positive cases or more were used to ensure the RF models had sufficient data to process (Table 2.2).

**Table 2.2.** Specific Positive and Negative Counts of Zoonotic Diseases Used in Random Forests Analysis

| No. | Virus/Isolate Host/Country/Year/Strain | Abbreviation | Counts |  |
| --- | --- | --- | --- | --- |
|  |  |  | Negative | Positive |
| 1 | Menangle virus/S. domesticus/AUS/2001 | MENV | 847 | 49 |
| 2 | Bombali ebolavirus/M. condylurus/SLE/2016/Predict_SLAB000156 | BOMBV | 854 | 42 |
| 3 | Zaire ebolavirus/H.sapiens/COD/1976/ Yambuku-Mayina | EBOV | 853 | 43 |
| 4 | Bundibugyo ebolavirus/H. sapiens/UGA/2007 | BDBV | 851 | 45 |
| 5 | Sudan ebolavirus/H. sapiens/UGA/2000/Gulu-808892 | SUDV | 850 | 46 |
| 6 | Mojiang henipavirus/R. sladeni/CHN/2014/Tongguan1 | MOJV | 851 | 45 |
| 7 | Ghanaian bat henipavirus/E. helvum/GHA/2009/GH- | GHV | 849 | 47 |

The data was divided into a training set and validation set. This is done so that the model could be evaluated on samples that were not used to build or fine-tune the model, so that they provide an unbiased sense of model effectiveness. For this study, 70% of the data set was used for training while 30% was used to test and validate the model.

The synthetic minority over-sampling technique (SMOTE) (Chawla et al., 2002) was used to balance positivity and negativity since positivity rates were low. The *smote* function in R was used to do the over-sampling. It is a data sampling procedure that uses both up-sampling and down-sampling, (Kuhn & Johnson, 2016). It creates “synthetic” examples instead of over-sampling with replacement (Chawla et al., 2002). The minority class is over-sampled by taking each minority class sample and introducing synthetic examples along the line segments joining any and/or all of the k minority class nearest neighbors. Neighbors from the k nearest neighbors are randomly chosen. This was done after the train/test splitting of the data. The *perc.over* function within *smote* was set to “100” to keep a 1 to 1 proportion to balance the “negative” and “positive” classes. It uses over-sampling to create the number of extra cases needed from the minority class to balance both classes. 1000 trees were used because the linear combination of many independent learners reduces the variance of the overall ensemble relative to any individual learner in the ensemble.

The most important measures for classifying disease are accuracy, sensitivity, specificity, positive predictive values, negative predictive values, ROC, and AUC (Trevethan, 2017; Lantz, 2019). Accuracy specifically measures how often the model trained is correct, which is depicted by using the confusion matrix (Chen et al., 2020). Sensitivity measures the proportion of positive examples that were correctly classified (Lantz, 2019). Specificity measures the proportion of negative examples that were correctly classified. The positive predictive value is the proportion of positive examples that are truly positive. The negative predictive value is the proportion of negative examples that are truly negative. The ROC is commonly used to examine the tradeoff between the detection of true positives while avoiding the false positives. The AUC treats the ROC diagram as a two-dimensional square and measures the total area under the ROC curve. AUC scores are interpreted by the following: Outstanding=0.9 to 1.0, Excellent/Good=0.8 to 0.9, Acceptable/Fair=0.7 to 0.8, Poor=0.6 to 0.7, and No Discrimination=0.5 to 0.6. Below is a table (Table 3) showing the above RF performance measures.

The features (Figures 2.2) that were extracted from the RF models, were the permutation runs which permutes values of the outcome, which leaves correlation patterns between predictor variables untouched (Degenhardt et al., 2019). Permutation feature importance were used instead of mean decrease in gini index feature importances because these can be biased (Degenhardt et al., 2019).

**Figure 2.2.**
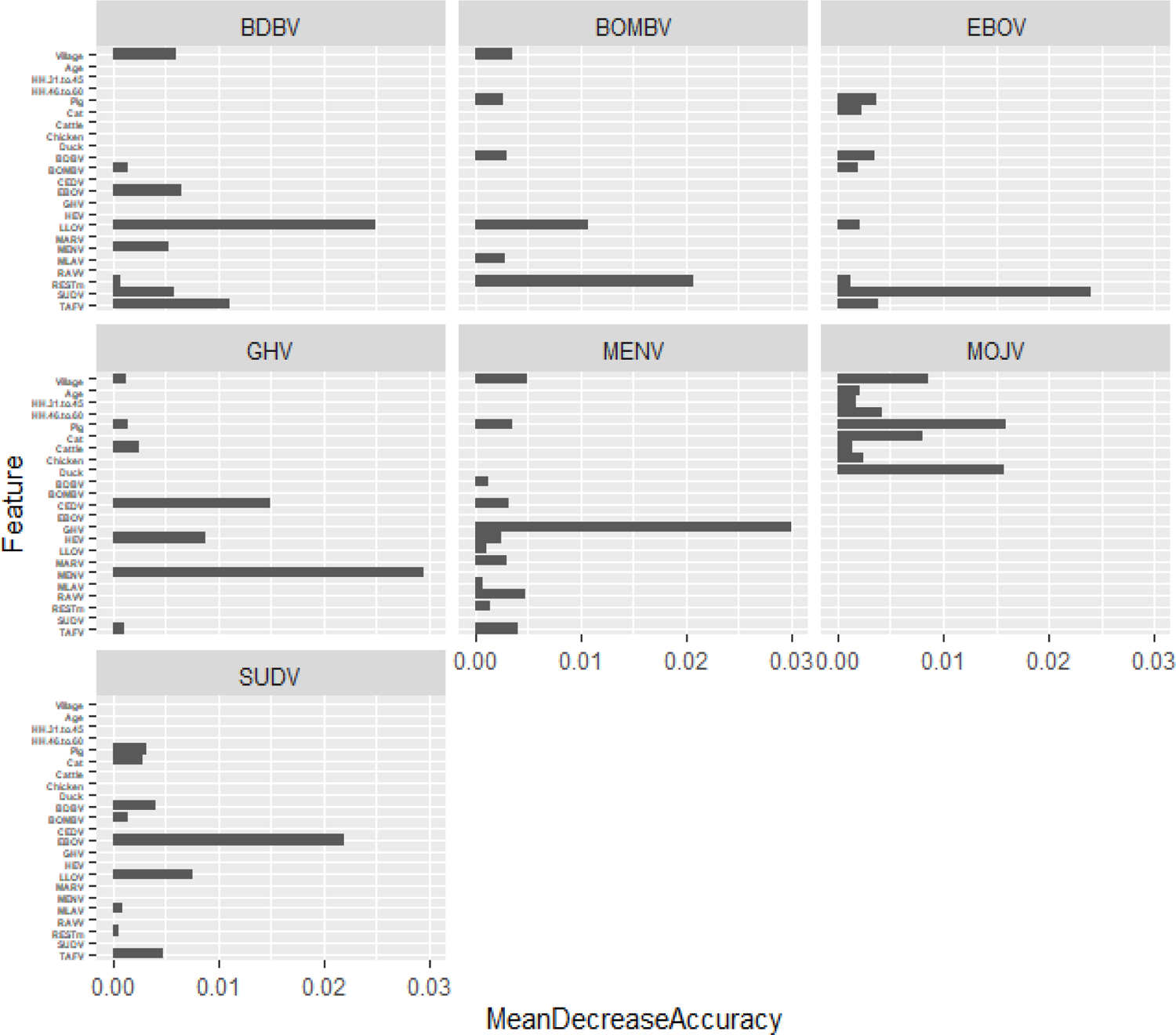
Random Forest Feature Importances For Each Disease

Before running the NB classifier, the data needs to be standardized so that the data are similar and so that the classifier can run properly (Géron, 2017). To standardize all the values, the *preprocessing* function, *scale*, and *center* were used to ensure all values were similar.

These functions are all part of the *caret* package created by Max Kuhn (2008). *Scale* divides the values by the standard deviation so that the dataset minimum is 0 and maximum is 1. *Center* was also used to subtract the mean from the values to scale the data to 0.

For the DT, scaling and centering were also done prior to the analysis using the *caret* package. The specific DT algorithm using the caret function was the *C5.0* algorithm. The C5.0 algorithm was used because it is the standard DT algorithm to use that works well across all types of data (Lantz, 2019). ANN was centered and scaled. The *caret* package in R was used to develop the ANN using the *nnet* function without any tuning. Nnet is a simple feed-forward ANN that uses a simple input layer, one hidden layer, and output layer.

All the algorithms above except RF were 10-fold cross validated. The out-of-bag error in RF is similar to cross validation especially if the classes are balanced (Janitza & Hornung, 2018).

## 3. Results

The RF model was able to accurately predict the proportion of true positives and true negatives divided by the total number of predictions via the accuracy score (Table 3.1). The lowest accuracy score is the RF model for MOJV (0.94). In positive disease classification, an important metric is sensitivity or the true positive rate. RF sensitivity scores for MENVV (0.67), BOMBV (0.57), EBOV (0.77), BDBV (0.64), SUDV (0.71), and GHV (0.67) were above 57%, but the MOJV, however, was 0.00.

**Table 3.1.** RF Classification Tree Statistics By Specific Disease

| **Disease** | **Accuracy** | **Sensitivity** | **Specificity** | **PPV** | **NPV** | **AUC** |
| --- | --- | --- | --- | --- | --- | --- |
| MENV | 0.96 | 0.67 | 0.98 | 0.67 | 0.98 | 0.92 |
| BOMBV | 0.96 | 0.57 | 0.98 | 0.62 | 0.98 | 0.88 |
| EBOV | 0.98 | 0.77 | 0.99 | 0.83 | 0.99 | 0.93 |
| BDBV | 0.97 | 0.64 | 0.99 | 0.82 | 0.98 | 0.95 |
| SUDV | 0.98 | 0.71 | 1.00 | 0.91 | 0.98 | 0.81 |
| MOJV | 0.94 | 0.00 | 1.00 | NaN | 0.94 | 0.75 |
| GHV | 0.96 | 0.67 | 0.98 | 0.67 | 0.98 | 0.88 |

The RF was able to correctly identify the proportion of negative cases that are truly negative via the negative predictive values (NPV). The RF model also had very high specificity scores (true negative rate). Additionally, according to the AUC scores, the RF models were able to distinguish between true positives (sensitivity) while avoiding false positives (specificity) except for MOJV which had a score of 0.75 (Figure 3.1).

**Figure 3.1.**
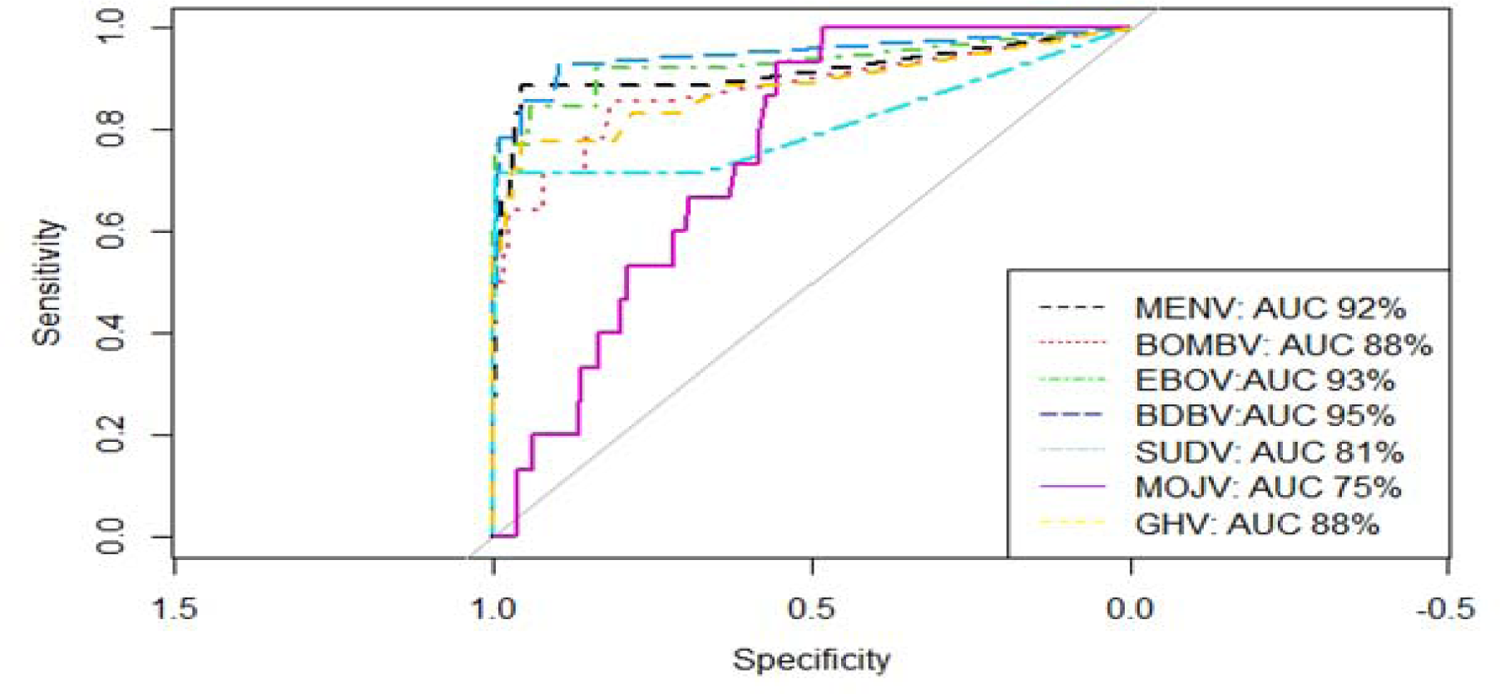
RF ROC graphs for each disease

The NB algorithm had the highest performance for each specific metric across all 7 viruses compared to RF, DT, and NN (Table 3.2). Compared to RF, NB has better performance when it comes to specificity, PPV, NPV, and ROC/AUC rates (Figure 3.2).

**Table 3.2.** Naïve Bayes Classification Statistics By Disease

| Disease | Accuracy | Sensitivity | Specificity | PPV | NPV | AUC |
| --- | --- | --- | --- | --- | --- | --- |
| MENV | 0.99 | 0.94 | 1.00 | 1.00 | 0.99 | 0.97 |
| BOMBV | 0.99 | 0.86 | 1.00 | 1.00 | 0.99 | 0.93 |
| EBOV | 1.00 | 1.00 | 1.00 | 1.00 | 1.00 | 1.00 |
| BDBV | 1.00 | 1.00 | 1.00 | 1.00 | 1.00 | 1.00 |
| SUDV | 1.00 | 1.00 | 1.00 | 1.00 | 1.00 | 1.00 |
| MOJV | 1.00 | 1.00 | 1.00 | 1.00 | 1.00 | 1.00 |
| GHV | 1.00 | 1.00 | 1.00 | 1.00 | 1.00 | 1.00 |

**Figure 3.2.**
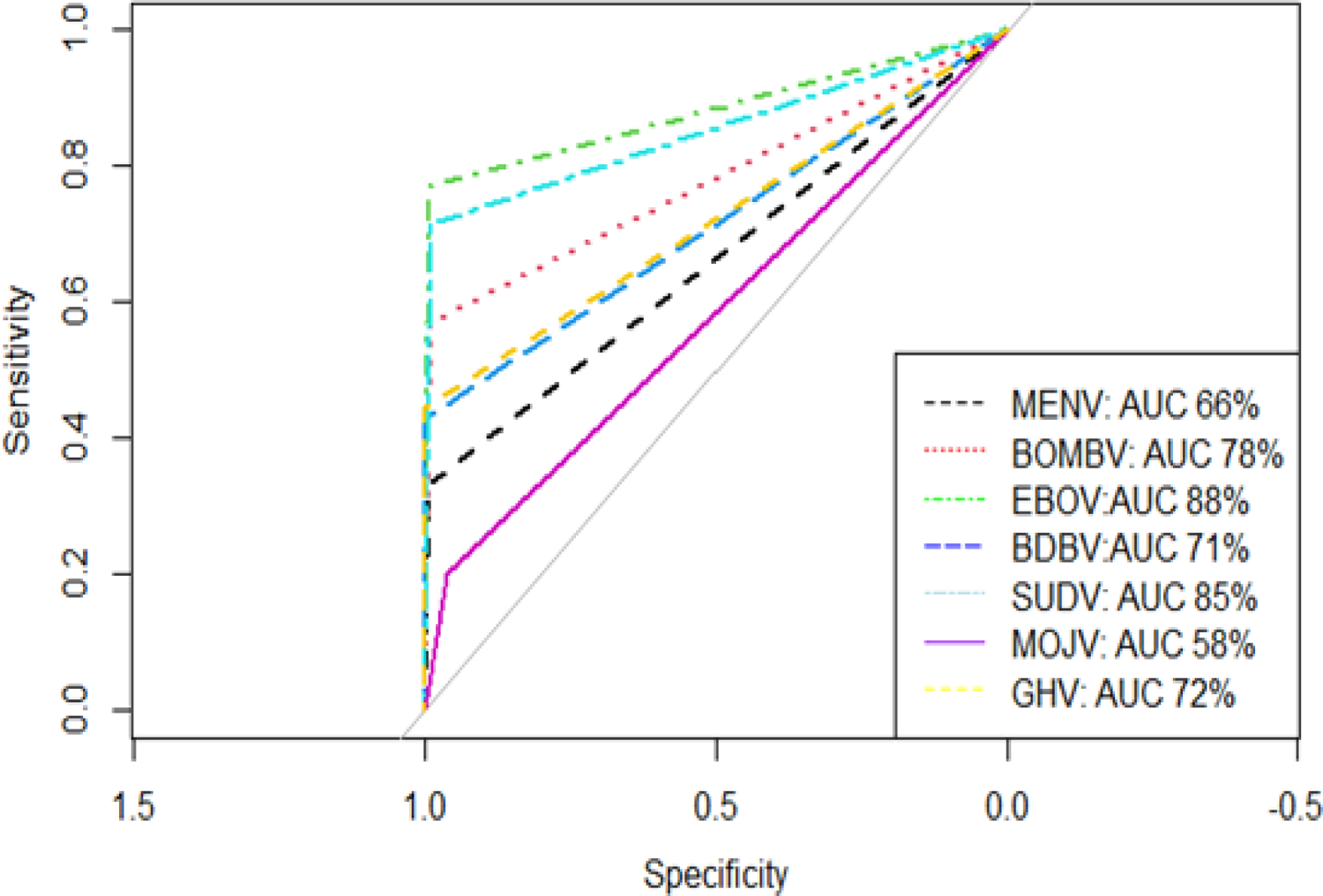
Decision Tree updated ROC graphs by specific disease

DT performed poorly and underfit the data which meant it was not able to capture all the data points and analyze it accordingly. Even though it works well in either small or large datasets, one of its weaknesses is that it is prone to underfitting (Lantz, 2019). For this study, it could not capture the data insights for MOJV (Table 3.3). Except for GHV, DT had poor performance regarding specificity and average results via the ROC/AUC scores for the rest of the diseases (Figure 3.3).

**Table 3.3.** Decision Tree Classification Statistics By Disease

| Disease | Accuracy | Sensitivity | Specificity | PPV | NPV | AUC |
| --- | --- | --- | --- | --- | --- | --- |
| MENV | 0.95 | 0.28 | 1.00 | 0.83 | 0.95 | 0.64 |
| BOMBV | 0.97 | 0.50 | 1.00 | 1.00 | 0.97 | 0.75 |
| EBOV | 0.99 | 0.77 | 1.00 | 0.91 | 0.99 | 0.88 |
| BDBV | 0.97 | 0.64 | 0.99 | 0.82 | 0.98 | 0.82 |
| SUDV | 0.97 | 0.71 | 0.99 | 0.77 | 0.98 | 0.85 |
| MOJV | 0.94 | 0.00 | 1.00 | NaN | 0.94 | 0.50 |
| GHV | 0.97 | 0.50 | 1.00 | 1.00 | 0.97 | 0.75 |

**Figure 3.3.**
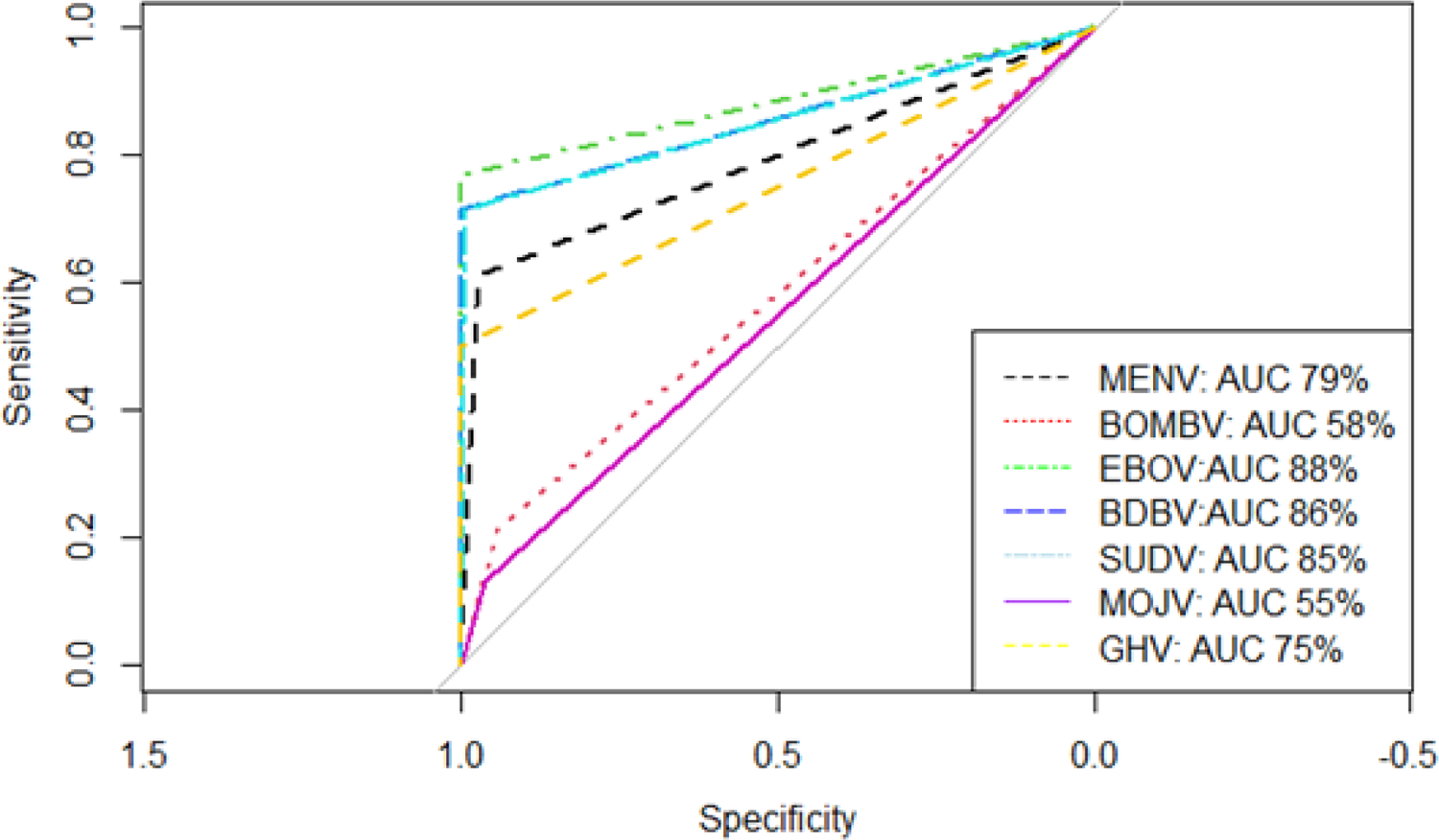
ANN updated ROC graphs by specific disease

The ANN algorithm captured the data and noise and thus, overfit the models for each disease (Table 3.4). Even though it only has one hidden layer, the ANN was still able to capture everything in the data by overfitting which it is prone to doing (Lantz, 2019). Thus, in the initial analysis, RF and NB classifiers had the best performance measure (Table 3.5). The ROC for all diseases using ANN was 1.00 (Figure 3.4).

**Table 3.4.** ANN Classification Statistics By Virus

| Disease | Accuracy | Sensitivity | Specificity | PPV | NPV | AUC |
| --- | --- | --- | --- | --- | --- | --- |
| MENV | 1.00 | 1.00 | 1.00 | 1.00 | 1.00 | 1.00 |
| BOMBV | 1.00 | 1.00 | 1.00 | 1.00 | 1.00 | 1.00 |
| EBOV | 1.00 | 1.00 | 1.00 | 1.00 | 1.00 | 1.00 |
| BDBV | 1.00 | 1.00 | 1.00 | 1.00 | 1.00 | 1.00 |
| SUDV | 1.00 | 1.00 | 1.00 | 1.00 | 1.00 | 1.00 |
| MOJV | 1.00 | 1.00 | 1.00 | 1.00 | 1.00 | 1.00 |
| GHV | 1.00 | 1.00 | 1.00 | 1.00 | 1.00 | 1.00 |

**Table 3.5.**
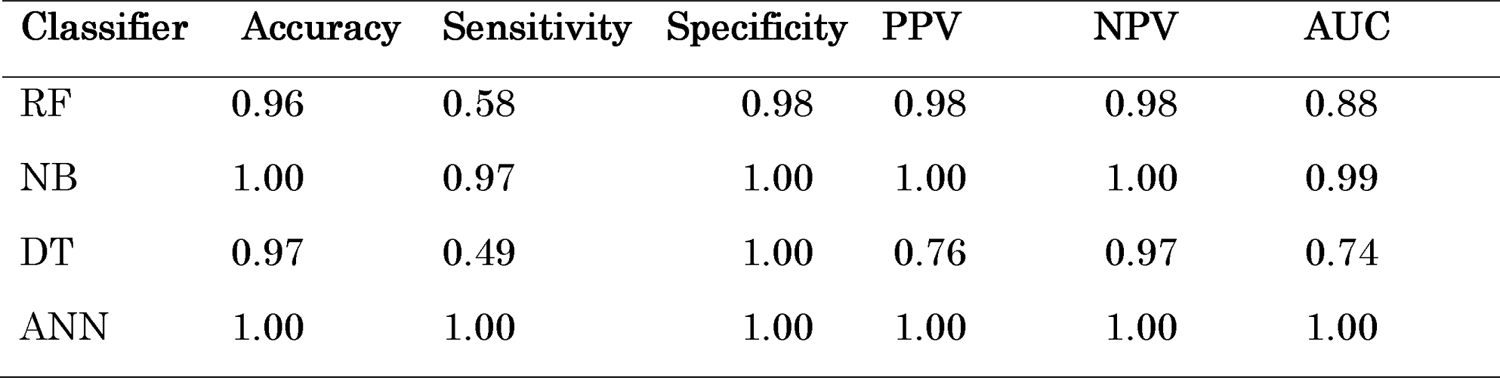
Initial Average Performance Comparison Between Random Forests, Naïve Bayes, Artificial Neural Network, and Decision Tree Classifiers

| Classifier | Accuracy | Sensitivity | Specificity | PPV | NPV | AUC |
| --- | --- | --- | --- | --- | --- | --- |
| RF | 0.96 | 0.58 | 0.98 | 0.98 | 0.98 | 0.88 |
| NB | 1.00 | 0.97 | 1.00 | 1.00 | 1.00 | 0.99 |
| DT | 0.97 | 0.49 | 1.00 | 0.76 | 0.97 | 0.74 |
| ANN | 1.00 | 1.00 | 1.00 | 1.00 | 1.00 | 1.00 |

To prevent underfitting, the DT method was changed to the *rpart2* method in the *caret* package was used instead of the C5.0 method used initially. *rpart2* uses maximum tree depth to have more nodes and splits and thus, captures more information and thus, it is better than the C5.0 algorithm. The *tunegrid* function was also used after the *rpart2* method was incorporated, but it did not change any of the performance measures. The *tunegrid* function finds the best performance using different combinations of parameters. rpart2 considerably captured more information via the ROC/AUC (Table 3.6, Figure 3.2) especially regarding EBOV which had average performance with the C5.0 algorithm. Nevertheless, compared with RF and NB, the sensitivity rates are very low.

**Table 3.6.**
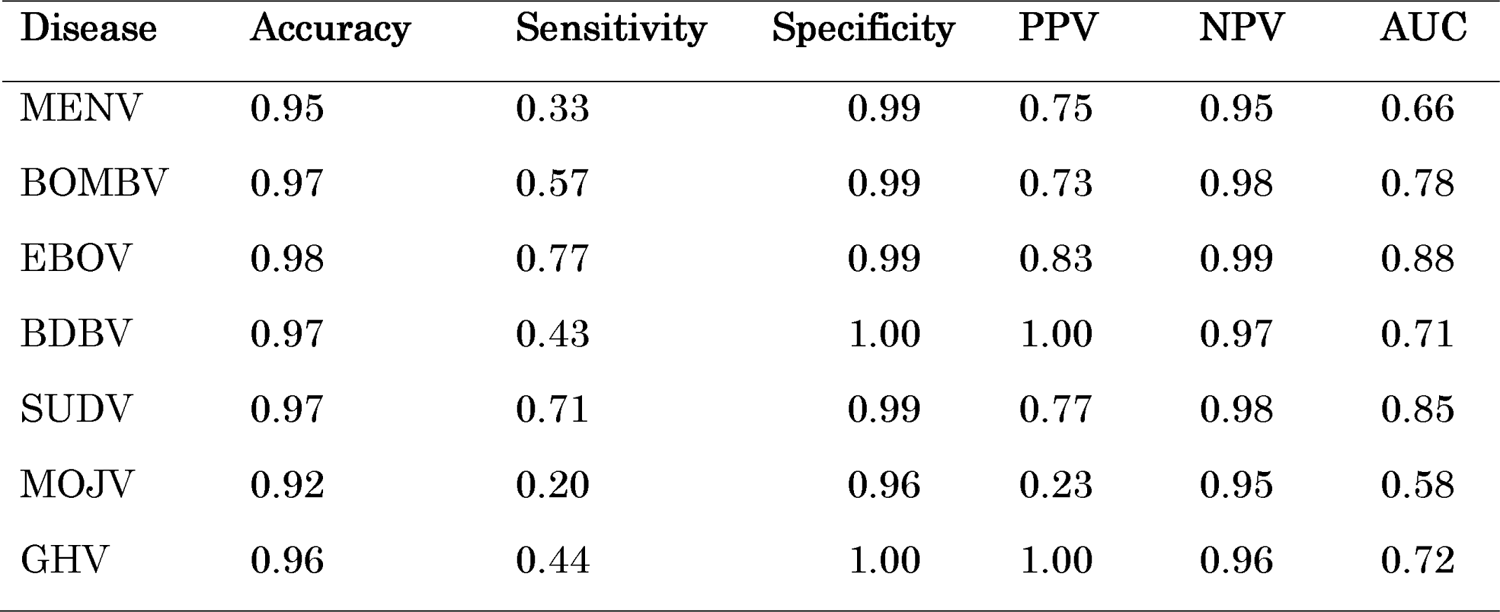
Decision Tree Updated Classification Statistics by Disease

| Disease | Accuracy | Sensitivity | Specificity | PPV | NPV | AUC |
| --- | --- | --- | --- | --- | --- | --- |
| MENV | 0.95 | 0.33 | 0.99 | 0.75 | 0.95 | 0.66 |
| BOMBV | 0.97 | 0.57 | 0.99 | 0.73 | 0.98 | 0.78 |
| EBOV | 0.98 | 0.77 | 0.99 | 0.83 | 0.99 | 0.88 |
| BDBV | 0.97 | 0.43 | 1.00 | 1.00 | 0.97 | 0.71 |
| SUDV | 0.97 | 0.71 | 0.99 | 0.77 | 0.98 | 0.85 |
| MOJV | 0.92 | 0.20 | 0.96 | 0.23 | 0.95 | 0.58 |
| GHV | 0.96 | 0.44 | 1.00 | 1.00 | 0.96 | 0.72 |

The *nnet* function was updated with the *tunegrid* function within the *caret* package to find the optimal parameters. Additionally, *size* and *decay* functions were added within the *tunegrid* to find the most optimal parameters. *Size* is the number of units in a hidden layer. For this update, it was set from 1 to 10 in increments of 1. The decay parameter is the weight decay regularization method used to prevent overfitting and it was set from 0.1 to 0.5 in increments of 0.1. This prevented the overfitting for most diseases and most performance metrics. However, it was not able to capture the data for the sensitivity rates for all the diseases (Table 3.7). Overall, the disease performance measures via specificity, PPV, and AUC do not compare to the RF and NB models which performed the best in this study (Table 3.7; Figure 3.3).

**Table 3.7.**
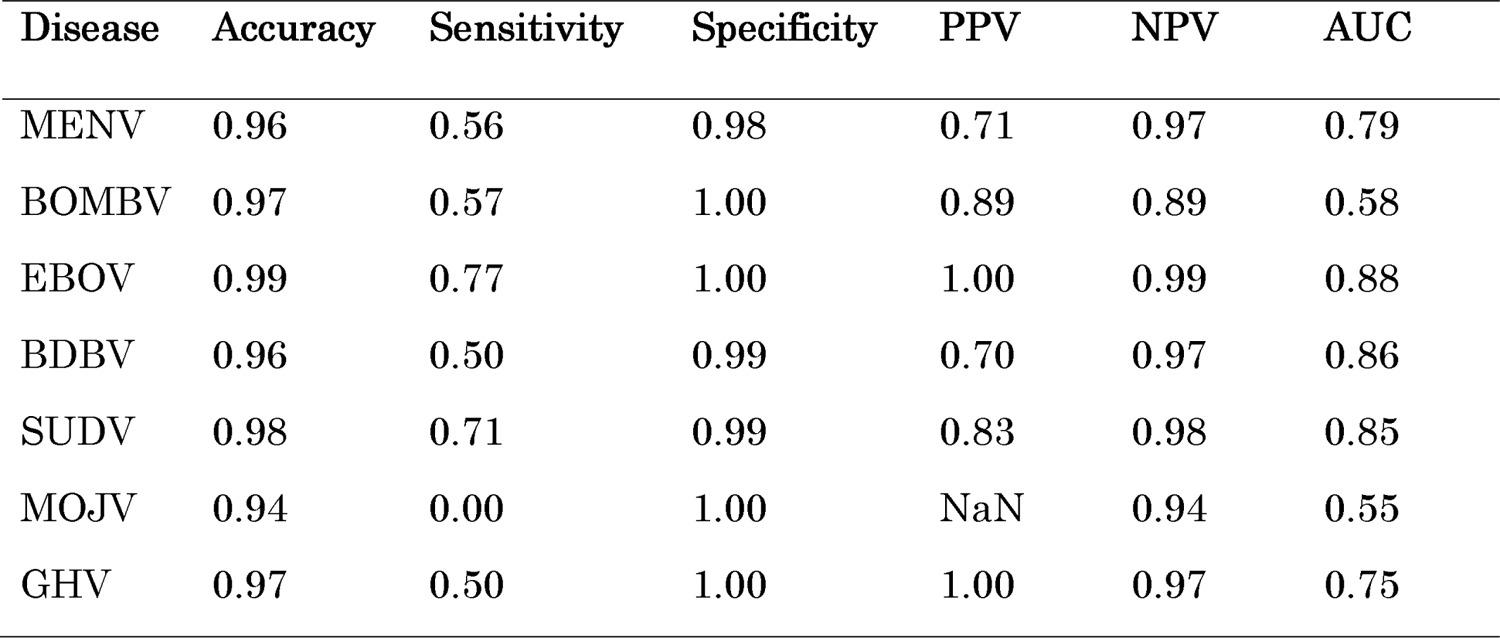
Artificial Neural Network Updated Classification Statistics By Disease

| Disease | Accuracy | Sensitivity | Specificity | PPV | NPV | AUC |
| --- | --- | --- | --- | --- | --- | --- |
| MENV | 0.96 | 0.56 | 0.98 | 0.71 | 0.97 | 0.79 |
| BOMBV | 0.97 | 0.57 | 1.00 | 0.89 | 0.89 | 0.58 |
| EBOV | 0.99 | 0.77 | 1.00 | 1.00 | 0.99 | 0.88 |
| BDBV | 0.96 | 0.50 | 0.99 | 0.70 | 0.97 | 0.86 |
| SUDV | 0.98 | 0.71 | 0.99 | 0.83 | 0.98 | 0.85 |
| MOJV | 0.94 | 0.00 | 1.00 | NaN | 0.94 | 0.55 |
| GHV | 0.97 | 0.50 | 1.00 | 1.00 | 0.97 | 0.75 |

## 4. Discussion

We tested four classifiers to predict infectious disease: RF, NB, DT, and ANN. Feature importances were extracted using the RF classifier. These feature importances were then used for the other classifiers. From an overall disease prevention perspective, the RF models above did well using PPV, AUC and ROC performance measures in identifying the positive cases for each of the diseases which is imperative when it comes to being able to identify the disease and then use this information to implement prevention and medical aid to specific areas and people where it is most needed (Table 3.8). It also does well in predicting the negative values which is important to ensure the negatives are not false negatives. RF works well because it is free from overfitting and outliers do not affect it (Byeon, 2020). It also generates high accuracy by reducing generalization errors.

**Table 3.8.** Average Performance Comparison Between Random Forests, Naïve Bayes, and updated Decision Tree and Artificial Neural Network Classifiers

| Classifier | Accuracy | Sensitivity | Specificity | PPV | NPV | AUC |
| --- | --- | --- | --- | --- | --- | --- |
| RF | 0.96 | 0.58 | 0.98 | 0.65 | 0.98 | 0.88 |
| NB | 1.00 | 0.97 | 1.00 | 1.00 | 1.00 | 0.99 |
| DT | 0.96 | 0.44 | 1.00 | 0.76 | 0.97 | 0.74 |
| ANN | 0.97 | 0.52 | 0.99 | 0.73 | 0.97 | 0.75 |

NB is the best choice for accuracy and performance (Table 3.8). NB model predictions are comparable to previous research showing how well NB works on various types of disease prediction including infectious disease prediction (Kamal Alsheref & Hassan Gomaa, 2019; Uddin et al., 2019; Fatima & Pasha, 2017). NB works well because it treats each feature as independent and thus, any change in one feature will not affect the other in the NB model (Latha & Jeeva, 2019).

Even though Uddin et al. (2019), Kamal Alsheref & Hassan Gomaa (2019), and Fatima & Pasha (2017) found that Decision Tree (DT) and Artificial Neural Network (ANN) algorithms were highly accurate machine learning classifiers for disease classification, these classifiers did not do well according to accuracy, sensitivity, specificity, positive predictive value, negative predictive value, ROC/AUC performance scores. DT could not capture the data (underfitting) during the first initial modeling and after hyper tuning. ANN captured all the data including noise (overfitting) in the initial model, but underfit after hyper tuning. Both DT and ANN are not recommended as classifiers for this dataset.

Previous KAP survey studies used linear and logistic regression which have limitations including assuming there is a linear relationship between independent variables and the dependent variable in a real-life infectious disease scenario which can be more complex. Another limitation to linear and logistic regression is that the independent variables must be known ahead of time and then fitted to specific models. In our study, RF was used to identify specific features (independent variables in linear and logistic regression) that were able to predict specific zoonotic diseases in the case of Cambodia. Our models did not have to be retrained to identify best-fitting models. However, we did have to hypertune our models to get the best performance from each machine learning classifier algorithm. Additionally, not all models performed well. Of the four different machine learning classifier algorithms we used, RF and NB had the best performance scores.

When it comes to infectious disease surveillance, RF feature extraction is important because it chooses the most important features that can be used in specific classifiers to predict infectious diseases. We showed that RF and NB had higher overall accuracies than DT and ANN. However, NB is the best choice because in this study for this data, it is the most accurate regarding measures for public health diseases which are important including accuracy, sensitivity, specificity, Positive Predictive Value, Negative Predictive Value, AUC, and ROC. Compared to previous research using machine learning classifiers to infectious disease prediction, our results were mixed since only RF and NB performed well in our study which is reflected in this previous research. DT and ANN did not perform well even though the previous literature says these perform well in a variety of disease prediction studies. This could be mainly due to our study being specific to Cambodia and the complexities of identifying the limited data that can predict zoonotic disease in this case.

## Data Availability

All data produced in the present work are contained in the manuscript.

